# Genome-Wide Association Study Identifies Protective Genetic Factors in Active Blood Donors Against Multiple Diseases

**DOI:** 10.1101/2024.06.18.24309089

**Authors:** Jonna Clancy, Jarkko Toivonen, Jouni Lauronen, Satu Koskela, Jukka Partanen, FinnGen, Mikko Arvas, Jarmo Ritari

## Abstract

The healthy donor effect (HDE) refers to the observed lower mortality rate among blood donors compared to the general population. While HDE arises due to healthier individuals being more likely to be able to donate, the extent to which it is influenced by genetic differences remains largely unclear. To elucidate the genetic basis of HDE, we conducted a genome-wide association study (GWAS) involving 53,688 active blood donors with extensive donation histories and 228,060 controls from biobank cohorts within the FinnGen project. Our results identified 2,973 genome-wide significant loci associated with several health-related endpoints and levels of proteins and laboratory values. The associated loci related not only to blood groups but also to predisposition to infections and somatic and mental diseases, suggesting that HDE genetics extends beyond blood donation eligibility criteria. In conclusion, in this study we show that HDE is partially explained by genetic factors affecting various disease categories.

## Introduction

Blood donors are a highly selected population due to the health status criteria they must meet to be eligible to donate blood. Along with a general survey of the donors’ current state of health, basic restrictions on blood donation suitability include serious illnesses such as severe cardiovascular conditions, cancer and epilepsy^1^. Some conditions that restrict blood donation suitability have a strong well-known genetic background, such as the association of *HLA* genes in type 1 diabetes (T1D)^2^ or in certain autoimmune diseases^3–5^. As a minimum hemoglobin level is required for donation eligibility, genetic variants affecting iron and hemoglobin levels (may) also result in donor selection. In addition to the blood donation eligibility criteria, the healthier lifestyle in general followed by the blood donors and self-selection caused by various factors, e.g. common cold, results in a known phenomenon, *healthy donor effect*, HDE, a form of membership bias, which can lead to detrimental effects in a research setting if not taken into account^6^.

Comprehensive selection of different blood groups, and especially a sufficient amount of emergency blood type O RhD neg suitable for most recipients, are basic requirements in blood banks. Due to the high need for emergency blood O RhD neg and strong immunogenicity of certain blood group antigens such as ABO, Rh and Kell^7–10^, donor recruitment is partially based on blood group genotype and results in enrichment of certain blood group antigens in a blood donor pool. Well-known associations between ABO blood group antigens and cardiovascular conditions exist, more precisely between red cell antigen O and reduced thromboembolic diseases^11,12^. Risk for bleeding disorder, von Willebrand disease, is known to be higher among individuals with red cell O antigen^13^. Pregnancy-induced hypertension has been previously described to occur more in RhD positive individuals^12^. Moreover, in hemochromatosis, blood donation diminishes the symptoms, and donors with hemochromatosis-associated gene variants are known to be enriched in the blood donor population^14^.

Blood donor samples have been used as a healthy control cohort in genetics and biomedical research due to their commitment and willingness to participate^15^. Voluntary and frequent blood donation offers unique opportunities for longitudinal sampling. In addition, blood donors exhibit positive attitudes toward the use of their samples and data in scientific research, if not directly required in patient care^16,17^. However, the enrichment of genetic variants with known disease associations or biological functions may result in unpredicted effects in biomedical research. To reveal the genetics of blood donation and to understand the possible genetic selection leading to the HDE, we conducted a genome-wide association study (GWAS) between active blood donors of the (Finnish Red Cross) Blood Service Biobank and FinnGen cohort (Figure 1).

**Figure 1.**
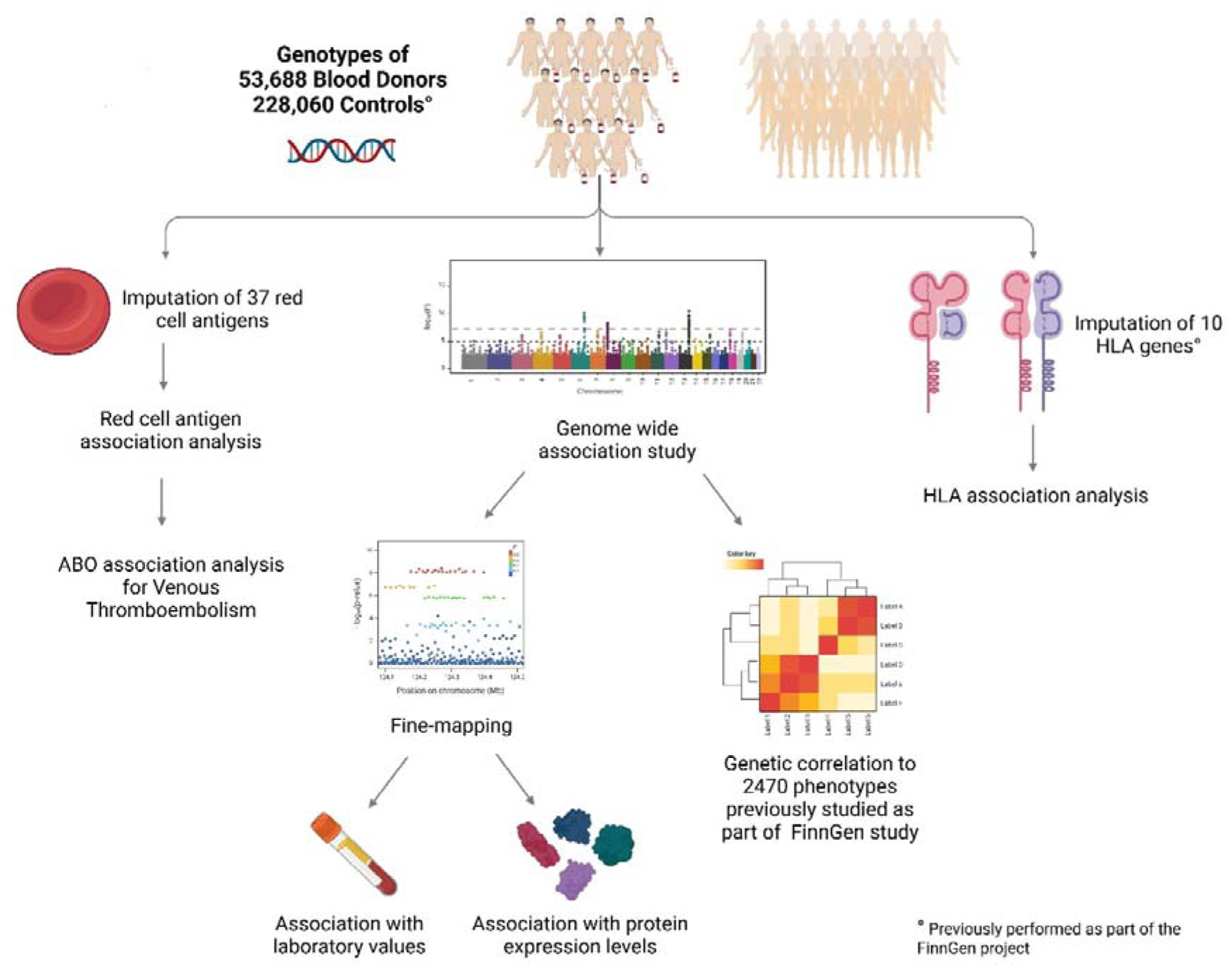
We conducted a GWAS on 53,688 blood donors and 228,060 controls (FinnGen). Genome-wide significant associations were fine-mapped and used for downstream analyses. Genetic correlations between 2,470 FinnGen phenotypes and blood donorship were calculated based on the GWAS summary statistics. Additionally, we imputed red cell antigens and tested their association with blood donorship. Previously imputed HLA alleles in FinnGen were similarly used for association analysis. We further used quantitative trait loci analysis to examine the association of fine-mapped and MHC lead variants with blood protein levels and clinical laboratory measurements. *Figure created with BioRender*.

To the best of our knowledge, no previous genome studies have focused on the possible selection bias caused by blood groups, health questionnaire-based requirement, and self-selection of blood donors. In our analysis, we found altogether 2,973 genome-wide significant associations in 46 distinct fine-mapped loci. Genetic correlation analysis revealed strong negative correlation between blood donorship and diseases such as mental disorders, pain disorders, cardiovascular diseases, addiction disorders, autoimmune disorders, and Alzheimer’s disease. *HLA* association analysis confirmed the enrichment of hemochromatosis-related and protective autoimmune *HLA* alleles and the lower occurrence of autoimmune-related *HLA* alleles in the blood donor population and indicated a risk SNP tagging both hemochromatosis and autoimmune related *HLA* alleles. The genome-wide significant variants also associated with 181 plasma protein level alterations and several clinical blood laboratory value measurements. The results of this study demonstrate the genetic basis of the HDE, which is not limited to immediate blood donorship inclusion criteria.

## Results

The overall study design is shown in Figure 1. Table 1 provides a detailed description of the study population characteristics and differences in diagnoses between the two cohorts. Blood donors, both male and female, were generally younger and taller than the controls. Additionally, male blood donors were heavier than their control counterparts. The blood donor cohort had a higher proportion of female participants. In all diagnosis categories, the prevalence of disease was significantly higher in the control group compared to the blood donors, except for the category ‘XVI Certain conditions originating in the perinatal period,’ where a higher prevalence of cases was observed among blood donors of both sexes. The FinnGen control cohort included donors from hospital biobanks, population cohorts, and occupational health settings (Supplementary Table 1).

**Table 1.** Characteristics of the study cohorts. Age, sex, height, weight, BMI and FinnGen endpoint categories in blood donors and FinnGen control cohorts are shown.

|  | Females |  | Males |  |
| --- | --- | --- | --- | --- |
|  | Blood donors | FinnGen | Blood donors | FinnGen |
| Median age at sampling, years (IQR) | 44.47 (18.2-70.8) | 62.7 (36.2-89.2) | 50.26 (25.5-75.0) | 70.46 (51.3-89.6) |
| Sex, n (%) | 32,460 (60.46) | 120,279 (52.74) | 21,228 (39.54) | 107,781 (47.26) |
| Median weight, kg (IQR) | 72 (54-90)<sup>a</sup> | 72 (51-93)<sup>a</sup> | 86 (68-104) | 84 (64-104) |
| Median height, cm (IQR) | 167 (159-175) | 164 (156-172) | 180 (171-189) | 177 (168-186) |
| Median BMI (IQR) | 25.83 (19.3-32.4) | 26.64 (19.1-34.2) | 26.58 (21.6-31.6) | 26.88 (21.2-32.6) |
| Number of diagnoses per FinnGen endpoint category (%)<sup>b</sup> |  |  |  |  |
| I Certain infectious and parasitic diseases (AB1) | 7,466 (23) | 47,901 (39.82) | 5,565 (26.22) | 48,487 (44.99) |
| II Neoplasms from cancer register (ICD-O-3) | 968 (2.98) | 34,299 (28.52) | 791 (3.73) | 38,891 (36.08) |
| II Neoplasms from hospital discharges (CD2) | 6,960 (21.44) | 67,026 (55.73) | 2,804 (13.21) | 52,708 (48.9) |
| III Blood and immune system (D3) | 964 (2.97) | 17,017 (14.15) | 534 (2.52) | 15,606 (14.48) |
| IV Endocrine, nutritional and metabolic (E4) | 10,269 (31.64) | 83,780 (69.65) | 3,029 (14.27) | 58,530 (54.3) |
| IX Circulatory system (I9) | 6,276 (19.33) | 69,429 (57.72) | 4,587 (21.61) | 78,876 (73.18) |
| V Mental and behavioural disorders (F5) | 7,871 (24.25) | 42,676 (35.48) | 3,648 (17.18) | 33,080 (30.69) |
| VI Nervous system (G6) | 6,431 (19.81) | 54,710 (45.49) | 4,501 (21.2) | 51,142 (47.45) |
| VII Eye and adnexa (H7) | 8,088 (24.92) | 58,567 (48.69) | 4,660 (21.95) | 47,174 (43.77) |
| VIII Ear and mastoid process (H8) | 5,316 (16.38) | 29,249 (24.32) | 3,893 (18.34) | 26,561 (24.64) |
| X Respiratory system (J10) | 14,408 (44.39) | 69,022 (57.38) | 11,532 (54.32) | 69,209 (64.21) |
| XI Digestive system (K11) | 27,542 (84.85) | 106,938 (88.91) | 17,037 (80.26) | 90,482 (83.95) |
| XII Skin and subcutaneous tissue (L12) | 7,710 (23.75) | 43,402 (36.08) | 4,096 (19.3) | 32,486 (30.14) |
| XIII Musculoskeletal and connective tissue (M13) | 13,715 (42.25) | 83,931 (69.78) | 9,118 (42.95) | 68,570 (63.62) |
| XIV Genitourinary system (N14) | 17,334 (53.4) | 91,665 (76.21) | 4,463 (21.02) | 49,729 (46.14) |
| XIX Injury, poisoning, external (ST19) | 15,936 (49.09) | 73,725 (61.29) | 13,051 (61.48) | 70,375 (65.29) |
| XV Pregnancy, childbirth and the puerperium (O15) | 20,560 (63.34) | 86,799 (72.16) | 0 (0) | 0 (0) |
| XVI Perinatal period (P16) | 535 (1.65) | 1,000 (0.83) | 263 (1.24) | 514 (0.48) |
| XVII Congenital malformations (Q17) | 2,018 (6.22) | 10,032 (8.34) | 1,231 (5.8) | 7,258 (6.73) |
| XVIII Abnormal clinical and laboratory findings (R18) | 16,800 (51.76) | 93,458 (77.7) | 9,125 (42.99) | 80,419 (74.61) |
| XX External causes of mortality (VWXY20) | 13,025 (40.13) | 64,874 (53.94) | 10,741 (50.6) | 62,034 (57.56) |
| XXI Contact with health services (Z21) | 24,581 (75.73) | 10,4068 (86.52) | 10,544 (49.67) | 72,915 (67.65) |
| XXII Codes for special purposes (U22) | 1,417 (4.37) | 5,906 (4.91) | 595 (2.8) | 4,310 (4) |

<sup>a</sup> Not significant
<sup>b</sup> Shortened for clarity

### Genome-wide association analysis

Genome-wide association analysis revealed 2,973 significant (p < 5×10^−8^) SNPs associated with blood donorship. After fine-mapping, 5 coding (4 missense and 1 inframe deletion) and 36 non-coding variants were detected (Figure 2A). In addition to these 41 variants, 5 non-coding lead variants in the extended MHC region (Figure 2B and C) were included in the downstream analyses (Table 2).

**Figure 2.**
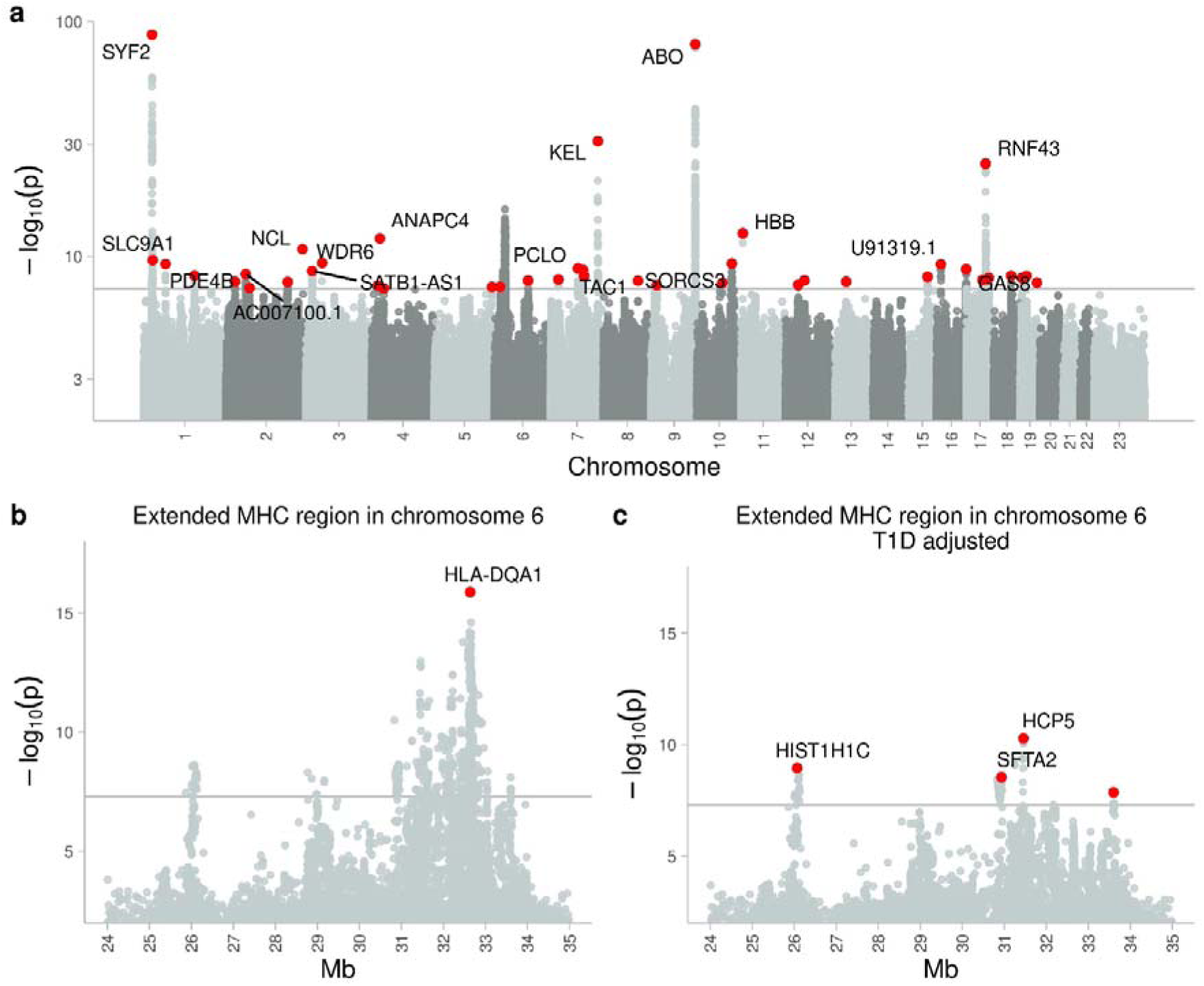
Results of the genome-wide association study (GWAS) for blood donorship. **a)** Manhattan plot of the whole genome. **b)** Extended major histocompatibility (MHC) region on chromosome 6. **c)** MHC association adjusted for type 1 diabetes (T1D). In panel **a)**, red dots indicate fine-mapped variants. In panels **b)** and **c)**, the highest variant in each peak reaching genome-wide significance (p < 5 × 10^−8^) is highlighted in red. For variants with p-values ≤ 5 × 10^−9^, the target gene or nearest gene is named. The horizontal line represents significance threshold of 5 × 10^−8^.

**Table 2.** Fine-mapped (41) and MHC lead variants (5) from blood donorship GWAS. P-value, effect size, the predicted most serious effect type and target gene for each variant are shown. Variants are ordered according to association effect size. In case the target gene is not known, the nearest gene is indicated by asterisk (*).

| chr | pos | Effect allele | rs number | pval | beta | Most severe | Gene most severe |
| --- | --- | --- | --- | --- | --- | --- | --- |
| 2 | 231460727 | T | rs66781921 | 1,89E-11 | -1,973 | inframe deletion | NCL |
| 11 | 5226799 | A | rs1135071 | 2,93E-13 | -0,573 | missense variant | HBB |
| 17 | 58358769 | T | rs199598395 | 1,55E-25 | -0,383 | missense variant | RNF43 |
| 7 | 142957921 | A | rs8176058 | 1,16E-31 | -0,327 | missense variant | KEL |
| 5 | 174443703 | C | rs191302298 | 3,89E-08 | -0,292 | intron variant | LINC01411 |
| 8 | 105038155 | C | rs10096658 | 1,35E-08 | -0,277 | intron variant | ZFPM2 |
| 19 | 5149612 | A | rs62113181 | 8,06E-09 | -0,135 | intron variant | KDM4B |
| 4 | 37135112 | C | rs75738358 | 4,89E-08 | -0,080 | intergenic variant | MIR4801* |
| 7 | 101559560 | A | rs12539059 | 5,68E-09 | -0,071 | downstream gene variant | COL26A1 |
| 6 | 32636375 | G | rs9272324 | 1,36E-16 | -0,064 | intron variant | HLA-DQA1* |
| 19 | 18707615 | AT | rs36035346 | 5,48E-09 | -0,062 | intron variant | CRTC1 |
| 7 | 23484043 | A | rs60678519 | 1,06E-08 | -0,055 | downstream gene variant | AC079780 , 1 |
| 6 | 30934404 | T | rs10947114 | 2,88E-09 | -0,055 | intron variant | SFTA2* |
| 6 | 31457404 | A | rs3130906 | 5,17E-11 | -0,055 | intron variant | HCP5* |
| 10 | 77157477 | A | rs80319585 | 1,92E-08 | -0,055 | intron variant | KCNMA1 |
| 7 | 82855734 | A | rs7795945 | 1,28E-09 | -0,049 | intron variant | PCLO |
| 2 | 186695046 | T | rs34483988 | 1,65E-08 | -0,049 | intron variant | FAM171B |
| 3 | 18717009 | T | rs6775319 | 2,17E-09 | -0,048 | intron variant | SATB1-AS1 |
| 10 | 105646256 | A | rs7907026 | 4,63E-10 | -0,048 | intergenic variant | SORCS3* |
| 1 | 153658595 | A | rs3806234 | 5,16E-09 | -0,047 | upstream gene variant | SNAPIN |
| 6 | 33602120 | C | rs4713637 | 1,39E-08 | -0,047 | intergenic variant | LINC00336* |
| 18 | 52600634 | C | rs62083414 | 5,35E-09 | -0,046 | intron variant | DCC |
| 4 | 19015485 | A | rs207350 | 3,03E-08 | -0,045 | intergenic variant | LCORL* |
| 17 | 48880334 | C | rs594398 | 1,18E-08 | -0,044 | intron variant | SUMO2P17 |
| 12 | 38272211 | T | rs10880819 | 2,64E-08 | 0,043 | downstream gene variant | RF00019 |
| 2 | 71295511 | CA | rs35440643 | 4,54E-08 | 0,044 | intron variant | ZNF638 |
| 2 | 26992548 | A | rs10166897 | 1,57E-08 | 0,044 | intron variant | MAPRE3 |
| 15 | 75059054 | TG | rs113236240 | 6,53E-09 | 0,045 | intron variant | PPCDC |
| 9 | 16695862 | CAAGCA | rs55763604 | 2,84E-08 | 0,049 | intron variant | BNC2 |
| 1 | 27087829 | T | rs55732343 | 2,31E-10 | 0,049 | regulatory region variant | SLC9A1* |
| 6 | 26071867 | C | rs9968910 | 1,12E-09 | 0,050 | intergenic variant | HIST1H1C* |
| 17 | 67991393 | T | rs5821444 | 7,98E-09 | 0,051 | 3 prime UTR variant | C17orf58 |
| 16 | 13464582 | C | rs6498415 | 5,47E-10 | 0,053 | intron variant | U91319 , 1 |
| 16 | 90023620 | G | rs12444334 | 1,51E-09 | 0,053 | intron variant | GAS8 |
| 2 | 59856883 | A | rs11675100 | 3,88E-09 | 0,054 | intron variant | AC007100 , 1 |
| 6 | 17127475 | A | rs9367942 | 3,81E-08 | 0,055 | intron variant | STMND1 |
| 12 | 57289173 | T | rs61937595 | 1,25E-08 | 0,062 | intron variant | R3HDM2 |
| 4 | 25407216 | A | rs34811474 | 1,21E-12 | 0,064 | missense variant | ANAPC4 |
| 1 | 66016472 | C | rs1500956 | 5,14E-10 | 0,075 | intron variant | PDE4B |
| 6 | 102401219 | C | rs77448558 | 1,28E-08 | 0,078 | intergenic variant | GRIK2* |
| 7 | 97662657 | T | rs10268629 | 1,71E-09 | 0,078 | intergenic variant | TAC1* |
| 13 | 50130043 | T | rs75350584 | 1,58E-08 | 0,094 | intron variant | DLEU1 |
| 9 | 133261662 | A | rs687621 | 5,70E-81 | 0,147 | intron variant | ABO |
| 1 | 25235176 | A | rs55794721 | 8,88E-89 | 0,157 | upstream gene variant | SYF2 |
| 19 | 50410043 | C | rs118101548 | 1,90E-08 | 0,299 | intron variant | POLD1 |
| 3 | 49008876 | T | rs528492111 | 4,09E-10 | 1,875 | intron variant | WDR6 |

The strongest genome-wide associations were seen in variants related to blood group antigens: rs55794721-A (p = 8.88×10^−89^) in chr 1, rs687621-A (p = 5.70×10^−81^) on chr 9 in *ABO* gene, and rs8176058-A (p = 1.16×10^−31^) on chr 7 in *KEL* gene (Figure 1A, Table 2).

Despite of the adjustment of the GWAS for BMI, we observed significant GWAS variants influencing height, weight, and BMI. rs66781921-T, an inframe deletion variant in chr 2, previously shown to have a negative association with height, weight and body mass index^18^, was rarer in blood donors (beta = −1.97, p = 1.89 × 10^−11^). The missense variant rs34811474-A, previously shown to be negatively associated with body mass index^18^, was found to be more frequent in blood donors (beta = 0.064, p = 1.21 × 10^−12^). Variants rs10947114-T (beta = −0.055, p = 2.88 × 10^−09^,), rs3130906-A (p = 5.17 × 10^−11^, beta = −0.055) and rs4713637-C (p = 1.39 × 10^−08^, beta = −0.047), previously described to be associated with spondylopathies and celiac disease^18^, were negatively associated with blood donorship. A rare intron variant in chromosome 3, rs528492111-T, previously shown to have a weak association to benign neoplasm of conjunctiva^18^, was more frequent in blood donors (beta = 1.87, p = 4.09 × 10^−10^). All the fine-mapped and MHC lead variants in the are listed in Table 2.

### Genetic correlation

To assess the genetic similarity between blood donors and various disease phenotypes, we performed a genetic correlation analysis comparing the blood donor GWAS with over 2,400 FinnGen core endpoint GWAS results.

Altogether 593 phenotypes were negatively correlated with blood donorship (FDR < 0.05, Supplementary File 2). Height and hemoglobin were the only phenotypes showing weak positive correlation with blood donorship, r_g_ = 0.07 and r_g_ = 0.18, respectively (Figure 3), despite that the blood donorship GWAS had been adjusted for BMI.

**Figure 3.**
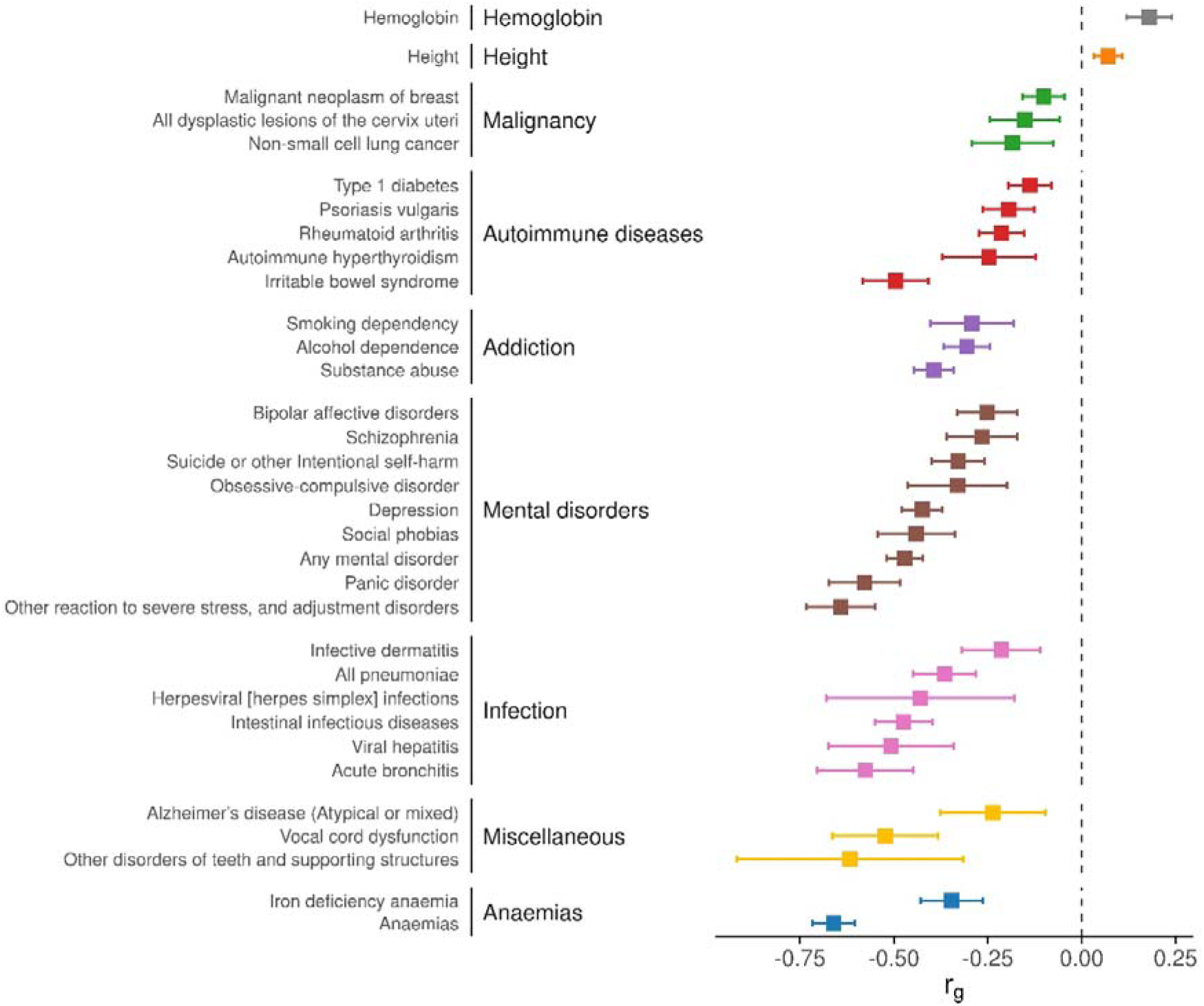
Genetic correlations of selected phenotypes against blood donorship phenotype. Each correlation is statistically significant at an FDR threshold of 0.05. The proportion of genetic variance shared between blood donorship, and a given phenotype (r_g_) is shown on the x-axis. Error bars represent the 95% confidence intervals. Apart from height and hemoglobin, all the phenotypes were negatively correlated with blood donorship.

Blood donorship was negatively correlated with r_g_ < −0.5 to 30 phenotypes (Supplementary File 2). The strongest negative correlation (r_g_ < −0.6) in blood donors was seen with ‘Anemias’ (r_g_ = −0.66, FDR = 2.81 × 10^−112^), ‘Other reaction to severe stress and adjustment disorder’ (r = −0.64, FDR = 3.36 × 10^−40^) and ‘Other disorders of teeth and supporting structure’ (r = −0.62, FDR = 2.74 × 10^−03^). Moderate negative correlation (r_g_ < −0.2) with malignancy-related phenotypes was observed. In addition, negative correlation with blood donorship was observed for addiction phenotypes such as smoking dependency (r_g_ = −0.29, FDR = 1.57 × 10^−05^), alcohol dependence (r_g_ = −0.31, FDR = 8.26 × 10^−20^) and substance abuse (r_g_ = −0.39, FDR = 2.88 × 10^−45^). Expectedly blood donors displayed protective correlation towards autoimmune related phenotypes. Protective correlation was also observed with autoimmune diseases in general (r_g_ = −0.33, FDR = 1.87 × 10^−52^), e.g. ‘Irritable bowel syndrome’ displayed strong negative correlation (r_g_ = −0.50, FDR = 4.05 × 10^−26^). A wide variety of mental illnesses (in total 49 phenotypes) were negatively correlated with blood donorship. The strongest mental illness-related phenotype with negative genetic correlation was ‘Mental disorders, not otherwise specified’ (rg = −0.59, FDR 0.001). Another example of phenotypes not directly restricted by blood donation eligibility criteria but with relatively strong negative correlation towards blood donorship were infectious phenotypes, such as acute bronchitis (r_g_ = −0.58, FDR = 1.76 × 10^−16^), viral hepatitis (r_g_ = −0.51, FDR = 1.88 × 10^−7^) and ‘other or unspecified bacterial infection’ (r_g_ = −0.50, FDR = 1.83 × 10^−09^) (Figure 3, Supplementary File 2). Lastly, phenotypes not directly prohibited by blood donation eligibility criteria, but which undeniably affect donation suitability or donor’s knowledge of their current state of health, such as Alzheimer’s disease (r_g_ = −0.24, FDR = 0.03), were also negatively correlated with blood donorship.

### Blood group antigen-related associations

To quantify the enrichment of blood group antigens among blood donors, we evaluated the association between imputed red cell antigens and blood donorship using a logistic regression model. The model revealed a strong negative association for the Kell, RhD, and AB blood group antigens, and a positive association for the O blood group antigen (Figure 4a). The effect sizes for the red cell antigens and other GWAS covariates are provided in Supplementary File 2.

**Figure 4.**
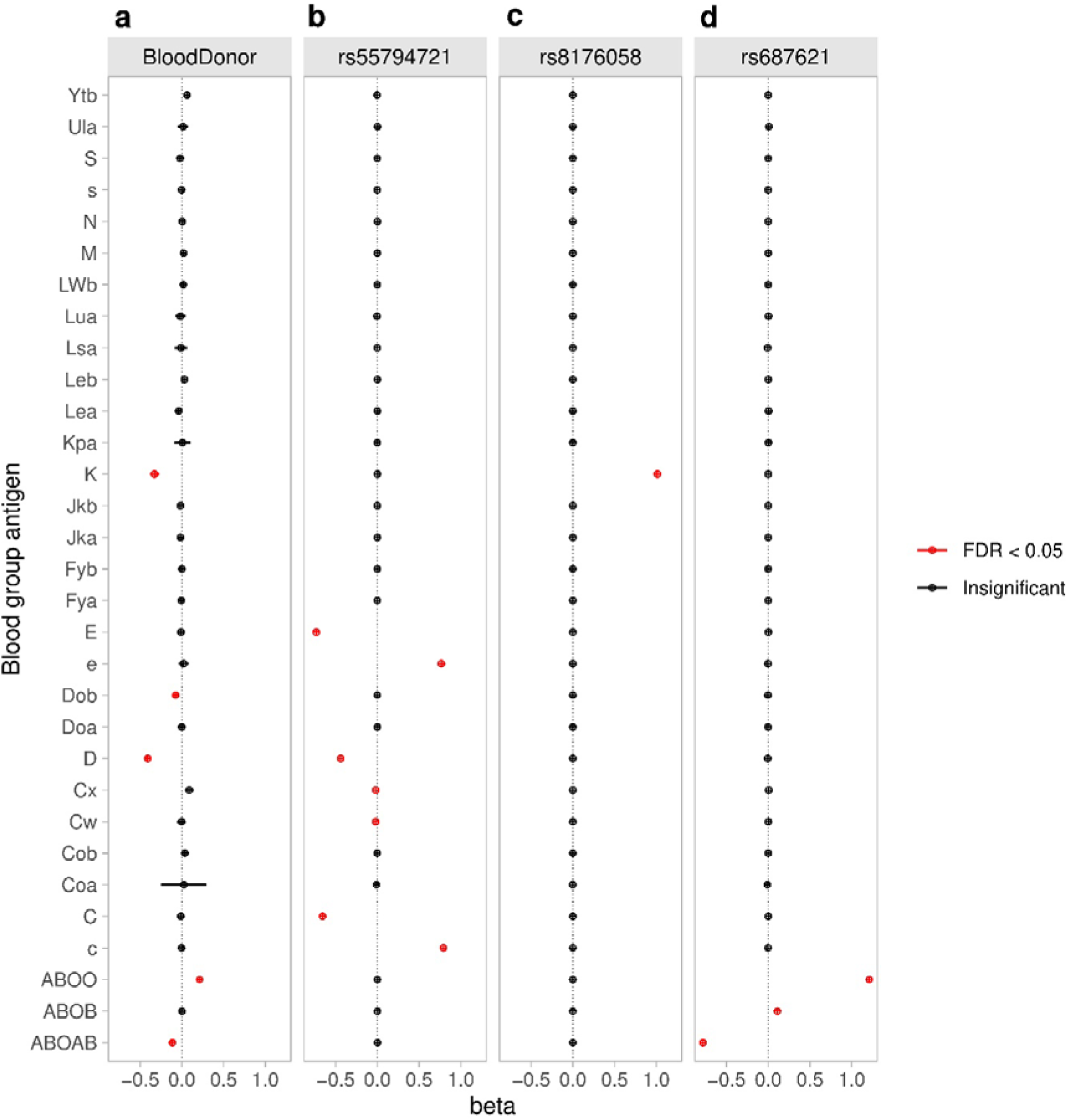
Association of blood group antigens with **a)** blood donorship and the fine-mapped lead variants **b)** rs55794721-A, **c)** rs8176058-A, and **d)** rs687621-A on chromosomes 1, 7, and 9, respectively. The magnitude of the association is represented on the x-axis as log odds ratios for the logistic regression of blood donorship and regression coefficients for the linear regression of the variant dosage.

We further analyzed the blood group related significant fine-mapped GWAS variants in chromosomes 1, 7 and 9 with linear regression models. The lead variant in chromosome 1, rs55794721-A, was associated with the Rh blood group system (previously shown to be associated with height^18^). Rh antigens E, C and D were negatively associated with rs55794721-A, whereas e and c antigens of the most common Rh negative haplotype in the Caucasoid population, dce^8^, were positively associated with rs55794721-A (Figure 4b). 91% of individuals positive for Rh c and e alleles and had no Rh D, C or E alleles, were homozygotes (dosage value > 1.5) for rs55794721-A. rs55794721-A is in LD^19^ (R^2^ = 0.39) with variant rs586178-C tagging certain RhCE blood group alleles^20^. Lead variant in chromosome 7, rs8176058-A, a known tag SNP for Kell blood group antigen^20^ was positively associated with K antigen (Figure 4c). The lead variant in chromosome 9, rs687621-A^19^, is in strong LD, R^2^ = 1 with blood group O tagging SNP rs687289-G^21^. We saw a strong positive association of rs687621-A with blood group antigen O, a positive association with blood group antigen B and a negative association with blood group antigen AB (Figure 4d).

### *HLA* allele associations

Because of the known strong role of the *HLA* region in immunology and the predisposing risk of certain *HLA* alleles in autoimmunity^2–5,22^, we performed an association study with imputed *HLA* alleles for blood donor endpoint. *HLA* association analysis for blood donorship (Figure 5, upper panel) revealed a negative association with *HLA*-alleles in linkage with the well-known T1D risk genotype *HLA-DQ2/DQ8*. A known risk allele for ankylosing spondylitis^22^ *HLA-B*27:05* was negatively associated with blood donorship. Positive association was seen with HLA alleles previously shown to be associated with HFE C282Y allele in Finland^14^ or HLA class II alleles with known protection from T1D^23^. To fine-map the MHC GWAS signal (Figure 2b), we adjusted the analysis with the MHC lead variant on chromosome 6 in *HLA-DQA1*, rs9272324-G (Figure 5, middle panel). We found that controlling for rs9272324-G decreased the effect sizes of the above-mentioned *HLA* alleles in linkage with *HLA-DQ8* and *HLA-B*27:05* and increased the p-values of the above mentioned HFE C282Y related *HLA*-alleles below the significance level. When adjusted for the lead *HLA*-allele, *DRB1*04:01* (Figure 5, lower panel), *HLA-B*27:05* and *HLA-C*02:02* in LD (r^2^ = 0.3) with *HLA-B*27:05* and HLA-alleles in linkage with HLA-DQ2 were negatively associated with blood donorship. Adjustment with *HLA-DRB1* revealed also the negative association of *HLA-B*08:01*, a known *HLA-B* allele in linkage with *HLA-DQ2* in Finland, with blood donorship (Figure 5, lower panel). In *HLA-DRB1*04:01* adjusted analysis, HFE C282Y-related *HLA* alleles *B*07:02*, *DRB1*15:01*, *DQA1*01:02* and *DQB1*06:02* were positively associated with blood donorship, but with less significant p-values than in *HLA*-association analysis without any additional adjustment. The most significant results of the HLA association analysis are shown in Supplementary Table 2 and full list of results in Supplementary File 2.

**Figure 5.**
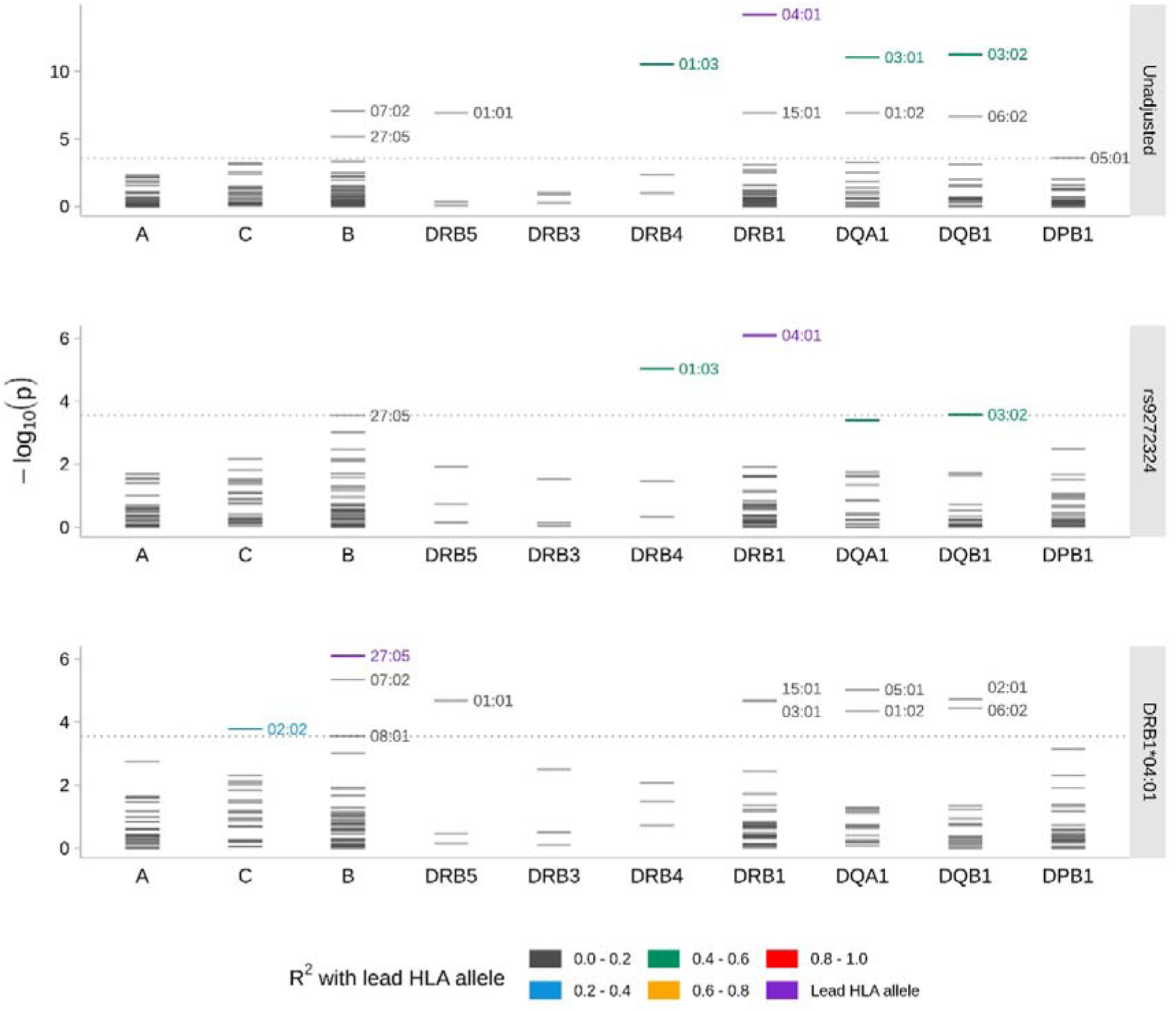
Conditional *HLA* association analysis. Upper panel: *HLA* association analysis for blood donorship. Middle panel: *HLA* association analysis for blood donorship adjusted for rs9272324-G. Lower panel: *HLA* association analysis for blood donorship adjusted for *HLA-DRB1*\*04:01. The other covariates used in all analyses were age, sex, BMI, PC1-10, birth region, and FinnGen genotyping array version. The dotted horizontal line represents the highest p-value that meets the FDR limit of 0.05.

### Association of the GWAS lead variants with molecular traits

To better understand the functional effects of the 46 selected GWAS lead variants, we used quantitative trait locus (QTL) analysis for protein expression levels, metabolites, and common electronic health care record-derived laboratory value measurements as well as functional enrichment analysis. Altogether 470,490 associations were tested, and 327 pQTL and labQTL associations reached the significance level of FDR < 0.05. Among the 41 fine-mapped and five MHC region variants, 19 were significantly associated with 184 unique plasma protein expression levels and 18 with 128 clinical laboratory value measurements (Table 3). However, no significant associations were observed between the 46 genetic variants and metabolite levels.

**Table 3.**
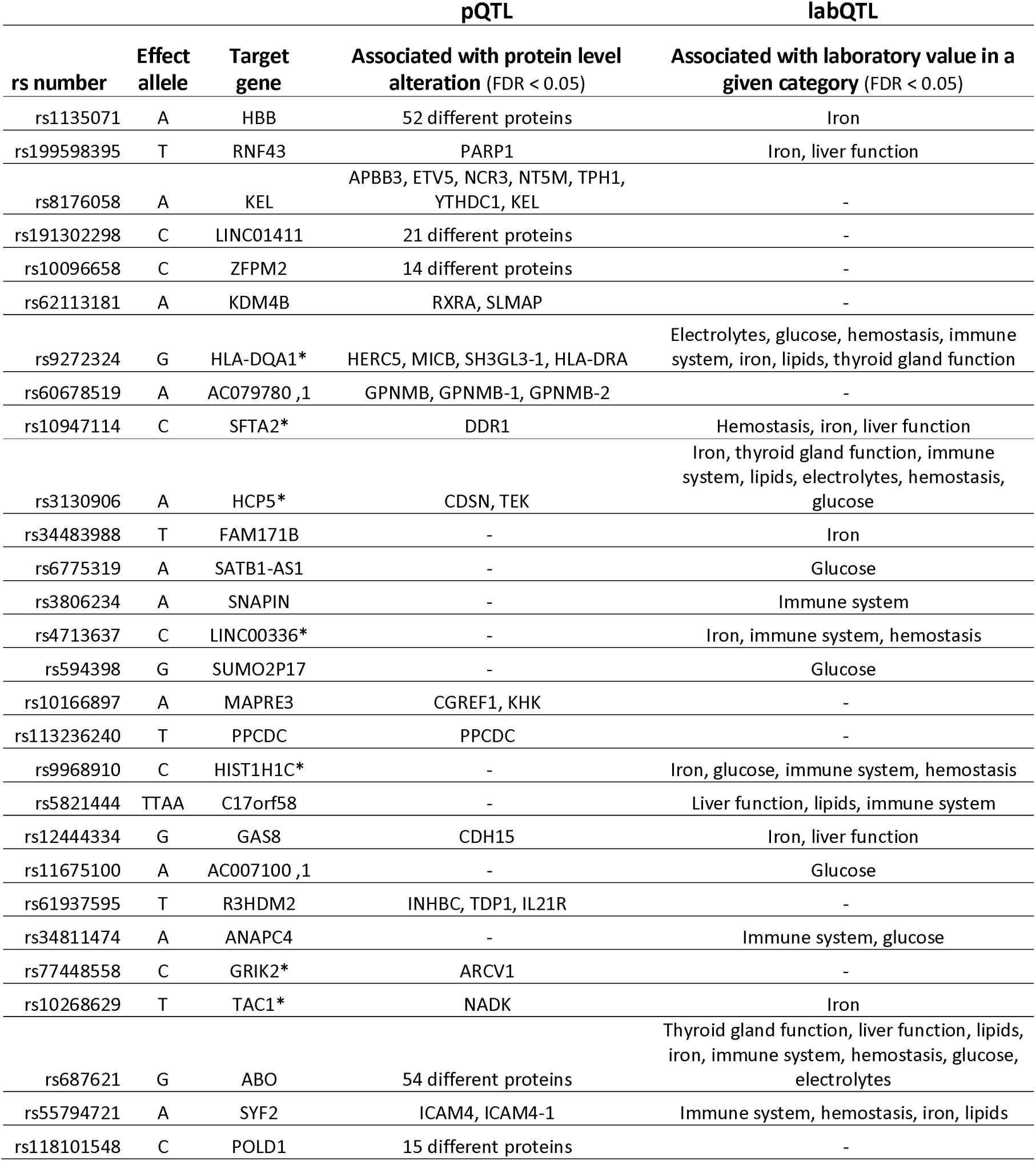
Significant associations (FDR < 0.05) of quantitative trait locus analysis for protein expression level and clinical laboratory values with the GWAS fine-mapped and MHC lead variants. In case the target gene is not known, the nearest gene is indicated by asterisk (*).

Since the proteomics data originated solely from the blood donor cohort, we were unable to analyze whether the significant pQTLs acted as mediators. In contrast, as the laboratory value measurement data comprised both blood donors and controls, we could perform a mediation analysis for the observed significant labQTLs. The analysis estimates the indirect effect of a genetic variant on blood donorship through a measured laboratory value.

#### Variants regulating protein expression levels

Two variants were shown to be associated with expression levels of more than 50 unique proteins (Supplementary Figure 2). rs-678621-G located on the *ABO* gene regulated expression levels of 54 proteins, and rs1135071-A in the *HBB* gene regulated 52 proteins. rs-678621-G pQTLs included proteins essential for the histo-blood group ABO formation, such as ABO itself and various N-acetylgalactosaminyltransferases such as GALNT9. Additionally, due to the enrichment of the rs687621-A allele in blood donors, decreased expression was observed for proteins such as F8 and MUC2, while the strongest increasing effect were seen for ALPI and SELE. Only two of the 52 associations identified with rs1135071-A showed decreasing effects on protein levels, the strongest effect was seen with TLR2, while the rest of the associations showed increasing effects, such as AHSP and MTIF3.

Other variants with more than 10 associations with protein expression levels included rs191302298-C (21 unique proteins), rs118101548-C (15 unique proteins) and rs10096658-C (14 unique proteins). In blood donors, the rs55794721-A variant exerted a decreasing effect on ICAM4, a LW glycoprotein present in the RhD protein complex. Additionally, a putative cancer-related protein association was observed for rs60678519-A and GPNMB. Due to the enrichment of rs60678519-AAAT in blood donors, GPNMB expression was decreased in blood donors. Overexpression of GPNMB has been linked to immunosuppression within the tumor immune microenvironment, leading to poorer prognosis in various cancers^24^.

The STRING^25^ functional enrichment analysis tool identified several Gene Ontology (GO) functional processes for the 184 proteins (Figure 6). Furthermore, the 54 proteins regulated by rs687621-G alone showed significant enrichments. Among the 184 proteins significantly associated with the 19 variants, GO revealed the enrichment of cell adhesion molecules in biological processes and signaling receptor activity in molecular functions. KEGG pathway enrichment was seen in cell adhesion molecules and mucin type O-glycan biosynthesis (Figure 6a, Supplementary file 3). Among the 54 proteins associated with rs687621-G, enrichment was seen in the biological process of cell adhesion and in molecular functions of antigen binding, transmembrane signaling receptor activity, carbohydrate binding, signaling receptor activity, transmembrane receptor protein tyrosine kinase activity and virion binding (Figure 6b).

**Figure 6.**
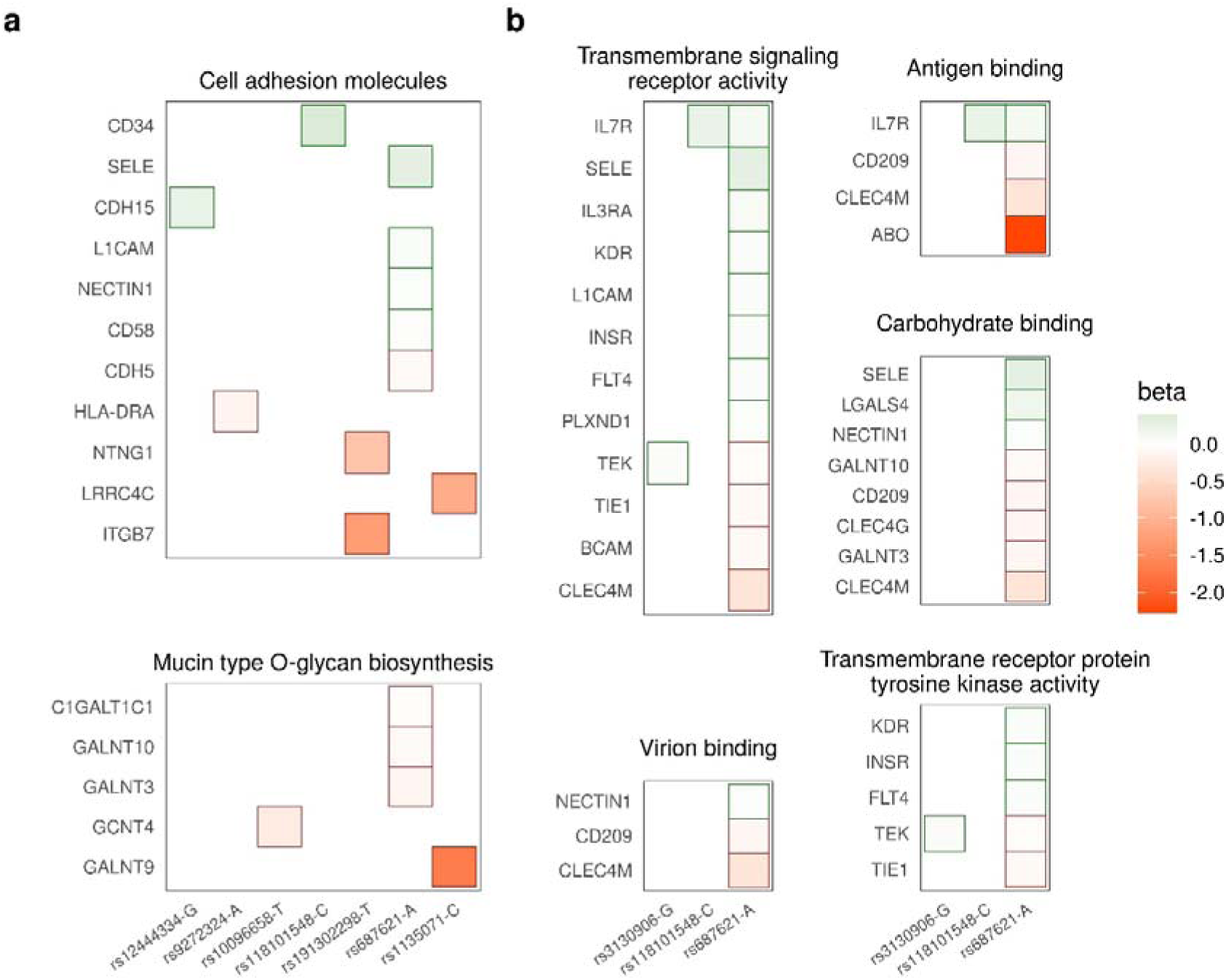
Enriched functional processes among the protein levels regulated by the 41 fine-mapped and 5 MHC lead variants. **a)** KEGG pathway enrichments among the 181 significant QTL target proteins. **b)** Five Gene Ontology (GO) functional processes enriched among the 54 proteins regulated by rs687621-A located in *ABO*. Three variants were found to regulate these functional processes. The associated variants and the effect alleles are shown on the x-axis and the proteins on the y-axis. The protein levels were measured in blood donors.

We detected 60 pQTLs that were not involved with the enriched biological processes or pathways. Of these, the strongest increasing and decreasing effect was seen with rs1135071-A on AHSP (beta = 2.59) and ACP6 (beta = −0.97), respectively (Supplementary Figure 2).

#### Variants regulating clinical laboratory test values

We observed 128 significant labQTLs that comprised nine variants and 50 different measurements and represented eight different clinical laboratory value categories (Supplementary File 4). Test of mediation for the top QTL of each 50 laboratory measurements showed significant mediation effect for 9 variants in 43 clinical laboratory measurements (Figure 7a). The highest number of mediated associations were seen in Iron (13 associations), Lipids (10 associations), Immune System (5 associations) and Liver Function (5 associations) related laboratory categories. The highest number of mediated associations, 16, on different laboratory values was seen for the MHC variant rs9272324. The results of the labQTL analysis are shown in detail in Supplementary File 4.

**Figure 7.**
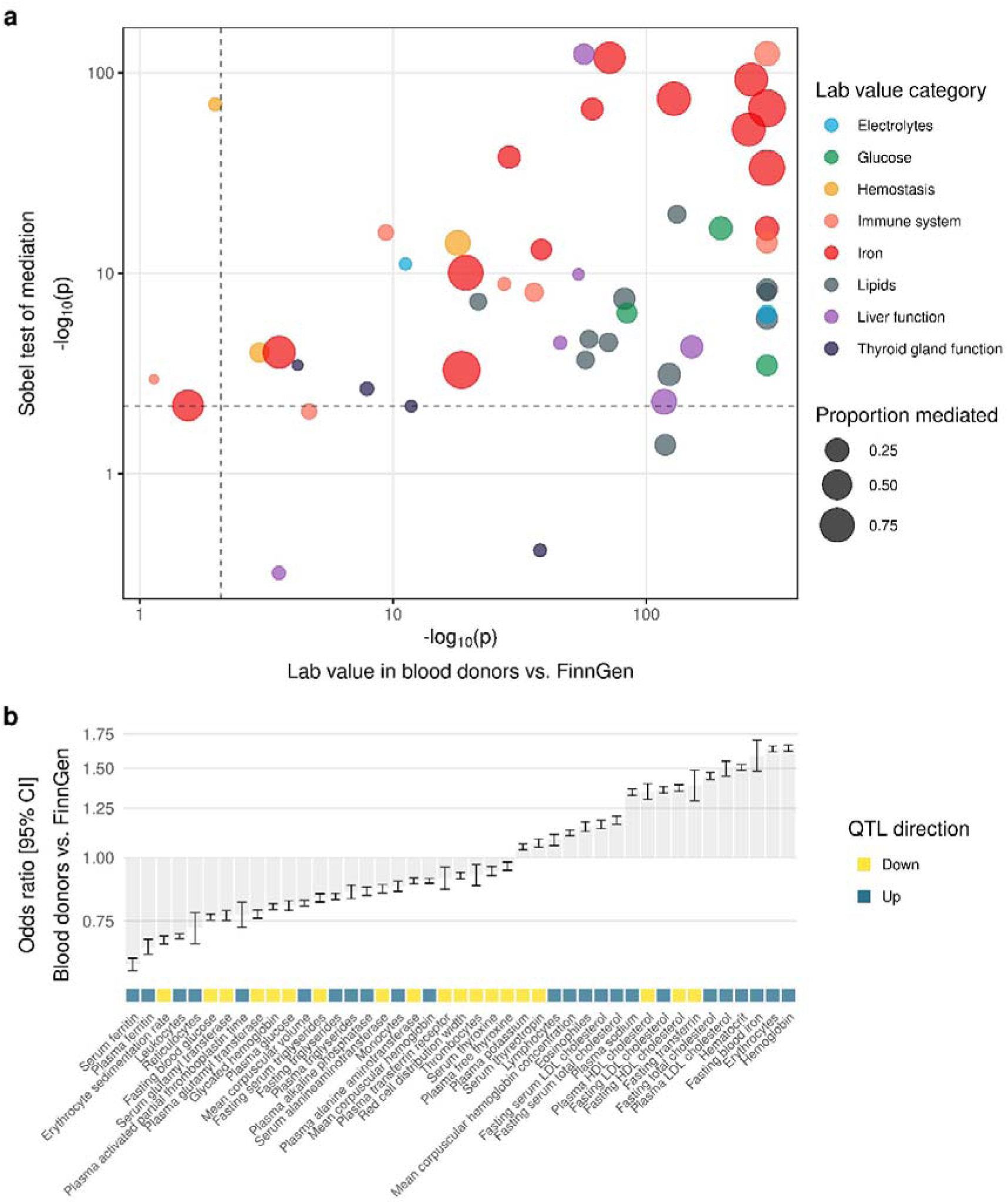
Mediation analysis of labQTLs. The analysis estimates the indirect effect of a genetic variant on blood donorship through a measured laboratory value. **a)** 43 significant mediation effects plotted in the top right quarter as defined by FDR < 0.05 difference in a lab value between blood donors and FinnGen controls adjusted for the QTL SNP (x-axis) and Sobel’s mediation test (y-axis). FDR limits are indicated by dashed lines. Proportion of mediation relative to the total effect of the variant is depicted by symbol size. **b)** Barplot of lab values with a significant QTL. The y-axis shows lab value effect sizes toward blood donorship, i.e. the effect size between blood donors and FinnGen controls when adjusted for the QTL SNP. The labQTL direction for the allele enriched in blood donors is depicted by the color bar on the x-axis. A lab measurement can have a lower value in blood donors than in controls (negative beta), but the QTL direction can still be up (or *vice versa*).

Strongest mediation effect, as measured by the proportion of mediated effect relative to the total SNP effect, was seen for iron related laboratory values. Glucose, hemostasis, lipids, immune and liver function related labQTLs also showed proportion of mediation > 0.2.

Of the 43 significant mediation effects, 25 showed lower and 18 higher clinical laboratory values in blood donors when compared to control population (Figure 7b). However, direction of the QTL effect on a laboratory value for the allele enriched in blood donors was sometimes opposite to the laboratory value difference between blood donors and controls. For example, blood donors had lower plasma ferritin levels (likely caused by donation), but they also harbored an enriched genetic variant that upregulates plasma ferritin.

### Putative bias caused by red cell antigen enrichment in research setting

Protective association of red cell antigen O to venous thromboembolism (VTE) has been shown previously by multiple studies^11–13^. To investigate the putative bias an ABO enrichment could cause to a VTE association study, we conducted an association analysis with VTE as an endpoint using chromosome 9 variants by adjusting for imputed ABO antigens. Our results showed that after the adjustment, the p-value of the lead variant failed to reach the genome-wide significance level of 5 × 10^−8^ (rs582118 unadjusted p = 3.28 × 10^−115^, ABO adjusted p = 1.16 × 10^−6^). The signal was not completely lost as other genome-wide significant variants remained in the region (Supplementary Figure 4A-B). The adjustment not only removed associations of some SNPs but also made visible other variants not detected in the basic analysis (Supplementary Figure 4C).

## Discussion

The healthy donor effect is a well-documented phenomenon, but its genetic basis has remained unclear. In this study, we conducted a genome-wide comparison between blood donor biobank donors with relatively long and active blood donation history^14^ and the FinnGen control population. The FinnGen controls included individuals from hospital biobanks, population cohorts, and patients at occupational health clinics. The latter two cohorts primarily comprise healthy individuals, and as disease-specific legacy cohorts were excluded from the analysis, the study controls are not ideal to but can be assumed to be not severely biased control population representing the general population of Finland.

The observed protection of blood donors from certain diseases can be directly attributed to donor eligibility criteria that select healthy individuals. However, some of the observed protective genetic factors may be secondary. For instance, the strongest negative genetic correlation was seen with phenotype ‘Anemias’. This correlation could be due to individuals with a higher tendency for diseases causing iron loss being less likely to start donating blood or to withstand repeated donations. Additionally, it may be a secondary result of autoimmune diseases, such as Crohn’s disease, which can restrict blood donation eligibility and lead to secondary anemia. Surprisingly, our results reveal that blood donors not only exhibit a protective genetic profile against diseases explicitly targeted by donor eligibility criteria, but also show a broader negative genetic correlation with conditions beyond these criteria, including various mental illnesses, infectious diseases and Alzheimer’s disease. This suggests that blood donors harbor a lower genetic disease burden than the general biobank population. While pleiotropic effects and/or self-selection may explain this phenomenon, our analyses indicate that the genetic correlation with the phenotypes targeted by the eligibility criteria is not consistently strong. Further detailed investigations are required to fully understand pleiotropy in blood donor selection and to explore the potential benefits of an active blood donor population for genetic and biomedical research.

Expectedly, genetic variants in the MHC region played strong role in several genetic associations and in QTLs. HLA alleles with known autoimmune associations occurred with lower frequency in the blood donor population than in the control population, whereas *HFE* C282Y-linked *HLA* alleles^14^ and *HLA* alleles with known protective role in autoimmunity were more frequent among the blood donors. Furthermore, the negative selection for T1D in blood donors leading to lower frequency of HLA-DQ2/8 genotype, could explain some of the protective genetic correlation. HLA-alleles of this genotype, have known predisposing associations, including certain infectious diseases and use of antidepressants^5^. These findings show the simultaneous genetic selection in the MHC for hemochromatosis and against autoimmune conditions among blood donors, while the pleiotropic role of the associated HLA alleles could explain some of the protective genetic correlation shown in this study.

In addition to tagging certain autoimmune-related *HLA* alleles or haplotypes (e.g. T1D and ankylosing spondylitis), rs9272324-G was shown to enhance the protein levels of *HLA-DRA*, which encodes the nonpolymorphic *DR* alpha-chain^26^, essential for the stable expression of DR molecules on the cell surface. Considering this, the known associations of *HLA-DRB1*15:01* with multiple sclerosis^3,27^ and the lower frequency of the rs9272324-G in blood donors, further investigation of the pleiotropic role of rs9272324 in multiple sclerosis and in autoimmunity in general would be of interest.

Furthermore, blood group antigen-related variants rs55794721 and rs687621 were found to decrease the expression level of genes involved in the function or structure of the cognate blood group molecules. These molecules include e.g. the RhD accessory protein ICAM4, the Landsteiner-Wiener glycoprotein^8^, and GALNT9, a type of N-acetylgalactosaminyltransferase necessary in the conversion of the *H* antigen into blood type A in ABO biosynthesis^13,28–30^.

In addition to hemochromatosis, the results of the present study highlight genetic factors that may affect recipient matching or donation activity. We show an increasing expression level of AHSP protein due to *Hb Tacoma* variant rs1135071-A. AHSP plays a crucial role in hemoglobin assembly by stabilizing the αHb subunits and preventing the self-aggregation of excess αHb subunits prior the formation of HbA1 subunit^31^. Given the speculated role of AHSP as a genetic modifier in β-thalassemia^31,32^ and the described normal clinical phenotype of *Hb Tacoma* heterozygotes^33^, rs1135071 could be of further interest in β-thalassemia research. Since the rs1135071-A allele is less common in blood donors compared to controls and strongly downregulates Hb and methemoglobin, it may be useful to analyze whether blood donation is harmless for *Hb Tacoma* heterozygotes or whether the blood products are suitable for recipients with conditions such as sickle cell disease^34^.

We demonstrated that blood donors differ from controls in several clinical blood laboratory measurements, such as ferritin and lipoproteins, that are regulated by the 41 fine-mapped and 5 MHC lead variants. For example, consistently with low prevalence of diabetes, blood donors exhibit lower plasma glucose levels, partly due to genetics, but also have higher LDL levels due to enriched QTLs. Interestingly, HDL levels are higher in blood donors, despite the presence of a variant that typically lowers HDL. This could be attributed to the U-shaped relationship between HDL and overall health^35^, where extremely high HDL levels are harmful and thus less common among blood donors. Our mediation analysis also revealed that blood donors had lower ferritin and higher hemoglobin levels than controls. This finding is in line with our previous studies; donation activity is a major determinant of ferritin in donors^36^, yet iron deficiency anemia is less common in donors (0.5 %) than in the Finnish general population (2.6 %)^37^. Consistent with this result, blood donors exhibited an enrichment of a compensatory variant that upregulates ferritin levels.

In this study we present the genetic underpinnings of HDE and blood group-based selection in active blood donors. Our findings highlight the unique genetic makeup of this donor population and its impact on health and blood donation suitability. The results demonstrate the role of genetics in maintaining good health status and active blood donation career which is also connected to genetic variation associated with a good mental health. Furthermore, research using blood donors as a control group should account for this distinct genetic structure, even after donation ceases. Additionally, the relatively predominant enrichment of specific blood group antigens among blood donors warrants consideration in research as these antigens have several known disease associations. No unambiguous solution exists to control collider biases when conducting research within blood donors, but rather a case-by-case evaluation e.g. through directed acyclic graphs can be helpful when evaluating the possibility of collider biases^38–40^. Further investigations are necessary to fully elucidate the implications of these discoveries, but they underscore the genetic structure behind HDE and the potential of active blood donor cohorts for advancing biomedical genetic research.

## Material and Methods

### Ethics statement

Study subjects in FinnGen provided informed consent for biobank research, based on the Finnish Biobank Act. Alternatively, separate research cohorts, collected prior the Finnish Biobank Act came into effect (in September 2013) and start of FinnGen (August 2017), were collected based on study-specific consents and later transferred to the Finnish biobanks after approval by Fimea (Finnish Medicines Agency), the National Supervisory Authority for Welfare and Health. Recruitment protocols followed the biobank protocols approved by Fimea. The Coordinating Ethics Committee of the Hospital District of Helsinki and Uusimaa (HUS) statement number for the FinnGen study is Nr HUS/990/2017. Full list of permits numbers are listed in Supplementary information.

All the sample donors had given a written biobank consent. The use of the data in the present study is in accordance with the biobank consent and meets the requirements of the Finnish Biobank Act 688/2012. A formal biobank decision 004-2022 for the research project has been granted by the Finnish Red Cross Blood Service Biobank. The use of the FinnGen data has been accepted with FinnGen proposal F_2022_086.

### Data acquisition

Genome data used in the present study originated from samples that were genotyped in the FinnGen project with the FinnGen ThermoFisher Axiom custom array v1 or v2 and genome imputation version R12. Genotyping, quality control, and genome imputation protocols are described in detail in FinnGen Gitbook (https://finngen.gitbook.io/documentation/). In short, genotype calling was performed with GenCall or zCall algorithm for Illumina and AxiomGT1 algorithm for Affymetrix arrays. Prior the imputation, genotyped samples were pre-phased with Eagle 2.3.5 with the default parameters, except the number of conditioning haplotypes was set to 20,000. Genotype imputation was performed using the population-specific imputation reference panel SISu v4.2 with Beagle 4.1 (version 27Jan18.7e1).

In addition to genome and phenotype data, the following electronic health register data and supplementing data were utilized: KANTA clinical laboratory values, ICD (ICD9 and 10) code for type 1 diabetes, occurrence of venous thromboembolism, age, BMI, sex, region of birth and genetic principal components (PC) 1-10.

To minimize potential bias in downstream analyses, legacy samples (https://finngen.gitbook.io/documentation/), highly enriched in certain common diseases, were excluded from the study controls due to the known selection of healthy individuals as blood donors. Individuals under 18 years of age were removed, as 18 is the minimum age of blood donation. In addition, individuals born in regions in Finland that were ceded to Soviet Union were excluded due to age structure of the region and the maximum blood donation age of 70 years. As BMI might influence donor eligibility, individuals lacking BMI information were filtered out. After filtering, total number of FinnGen genome data used in this study was 53,688 cases (blood donors) and 228,060 controls (other FinnGen participants).

### Statistical analyses

If not stated otherwise, all statistical analyses were performed in R^41^ version 4.3 or 4.4, with RStudio^42^. The genome-wide significance p-value threshold p < 5 × 10^−8^ was used in GWAS. In all other analyses, p-values were adjusted with the Benjamini–Yekutieli procedure^43^, and significance threshold of 0.05 was used. All the phenotypes analyzed here have been described in FinnGen DF12 dataset. The FinnGen summary statistics data are publicly available (https://r12.finngen.fi/)^18^, and the methods are available at https://finngen.gitbook.io/documentation/methods/.

### Association analysis and fine-mapping

Genome-wide association study, GWAS, using an additive genetic model, was performed in FinnGen pipeline with Regenie^44^ (https://github.com/FINNGEN/regenie-pipelines) v2.2.4 with blood donor status as a binary endpoint. Sex, age, BMI, PC1-10, birth region, and FinnGen genotyping array version were used as covariates in the GWAS. To discover associations independent of type 1 diabetes (T1D) in chromosome 6, the FinnGen T1D endpoint^45^ was used as an additional covariate in the chromosome 6-specific association analysis.

Fine-mapping for the genome-wide significant GWAS signals (p < 5 × 10^−8^) was performed in FinnGen pipeline (https://github.com/FINNGEN/) with SuSiE^46^ excluding MHC region (GRCh38, chr6:25-34 Mb)^45,47^. For each fine-mapped locus we accepted the credible set as an independent hit if the strongest primary p-value was p < 5 × 10^−8^. In case multiple credible sets within a single locus were found, we required genome-wide significance and log_10_(Bayes factor) > 2. As the MHC region is excluded from the fine-mapping pipeline due to its strong LD, four MHC lead variants from the T1D adjusted association analysis were selected for downstream analyses: rs9968910, rs10947114, rs3130906 and rs4713637. Moreover, the lead variant in the MHC region in the non-T1D adjusted GWAS, rs9272324, was included in the further analyses due to its strong association with autoimmunity^18^.

Association analyses with the blood donor endpoint for all the covariates used in GWAS and the imputed blood groups were performed with logistic regression in R. We further analyzed the associations of all the covariates used in GWAS and the imputed blood group antigens with the lead variants in chromosomes 1 (rs55794721), 7 (rs8176058) and 9 (rs687621) by using linear regression.

Association testing of each classical *HLA* allele *HLA-A*, -*B*, -*C*, -*DRB1*, -*DQA1*, -*DQB1*, *DPB1* and *HLA-DRB3-5* with the blood donor endpoint was performed using Regenie v3.0.1. First, a sparse genotype data set for fitting Regenie null models (https://rgcgithub.github.io/regenie/options/) was generated with Plink2 v2.00a4LM using FinnGen DF12 HapMap3 variants as a basis, and randomly thinning the variants to approximately 500,000 with further filtering for minor allele frequency at least 0.1, Hardy-Weinberg equilibrium p-value > 1 × 10^−15^ and minimum allele count at least 100. Second, the Regenie null models were fitted on the filtered genotype data and used in the following association step. To dissect the HLA signal further, two additional association tests were performed: i) conditioning for the GWAS chromosome 6 lead variant rs9272324 and ii) conditioning for the HLA allele with the strongest association, *HLA-DRB1*04:01*, in the primary HLA association analysis.

Since both the GWAS lead variant rs687621 in chromosome 9 and the red cell antigen ABO have previously been reported to be strongly associated with venous thromboembolism^45^(VTE), we sought to study whether ABO enrichment in the blood donor pool and the lead variant associate independently with VTE. To this end, we conducted an association analysis of chromosome 9 in FinnGen pipeline using Regenie^44^ (https://github.com/FINNGEN/regenie-pipelines) v2.2.4 with FinnGen venous thromboembolism as the binary endpoint. Sex, age, PC1-10, FinnGen genotyping array version, and FinnGen batch information were used as covariates, as well as imputed ABO red cell antigens. After filtering out 20 individuals based on low posterior probability of the imputed ABO antigens, 19,956 cases and 344,653 controls remained in the conditional analysis.

### Genetic correlation

All pair-wise genetic correlations between blood donorship and all the 2470 phenotypes previously studied as part of FinnGen study were computed using LD Score Regression v1.0.1 using Finnish LD panel in FinnGen pipeline (https://github.com/FINNGEN/)^48^. The computation was based on the GWAS summary statistics previously obtained using Regenie for each phenotype. We used the Benjamini-Yekutieli method to control FDR of genetic correlation at the level of 0.05. Genetic correlation between groups of phenotypes was approximated as the average of all the pair-wise genetic correlations between the groups.

### HLA and blood group imputation

The alleles of the classical HLA genes, *HLA-A*, *-C*, *-B*, *-DRB1*, *DRB3-5*, *-DQA1*, *-DQB1*, and *-DPB1*, were previously imputed at four-field resolution (defining protein sequence variation) in FinnGen using HIBAG^49^ algorithm with population-specific reference panel, as reported earlier^50^. The dosage value of each HLA-allele was used in downstream analyses.

Altogether 37 red blood cell antigens in 14 different blood group systems were imputed using population-specific random forest models as described by Hyvärinen et al^51^. *P1*, *A1* and *A2* antigens were excluded from downstream analyses due to uncertain frequency at the population level. In addition, blood group antigens *HrS* and *HrB*, occurring mainly in African population, are in high correlation with the blood group antigen *e* and have low clinical significance (consultation with an expert at the Finnish Red Cross Blood Service), hence they were excluded from further analyses. After imputation, the samples were categorized based on the posterior probability, PP, value; samples with PP < 0.5 were considered as antigen negative and samples with PP ≥ 0.5 were considered as antigen positive. Altogether 13 individuals were excluded from downstream blood group related analyses due to all PP values in ABO being < 0.5.

### QTL analysis

Proteomics data were previously generated in FinnGen on blood donors using multiplex antibody-based immunoassay (Olink) and multiplex aptamer-based immunoassay (SomaScan)^52^. We analyzed protein QTLs by performing an association analysis between 46 GWAS lead variants and protein expression data using glm function of Plink2 v2.00a4LM. Age, sex, sampling year and PC1-10 were used as covariates in the association analysis, and first-degree relatives were removed. To evaluate the enrichment of the associated proteins in functional processes, we used STRING interaction network and functional enrichment analysis tool^25^.

We also analyzed metabolite level data produced by Nightingale NMR platform (https://research.nightingalehealth.com/) in FinnGen. The data were preprocessed by selecting variables that were common between all available data files, removing duplicated sample IDs, and removing variables with >500 missing entries. The association analysis between cleaned NMR data and lead GWAS variants was performed as described above for proteomics.

The KANTA clinical laboratory test data in FinnGen spans from 2014 to 2023, as detailed in the FinnGen documentation (https://finngen.gitbook.io/finngen-handbook). Laboratory values were extracted for each individual in this study, comprising 53,688 blood donors and 228,060 controls. Missing values were excluded, retaining measurements with n > 100,000. The data were then cleaned by removing measurements > 5 standard deviations from the population mean. To obtain a single value per variable for each individual, we computed the geometric mean over time. To account for temporal changes in lab values, we also calculated the slope for each variable and individual. Finally, each variable was transformed into a standard normal distribution using quantile normalization.

We tested the associations of the 46 GWAS lead variants with the laboratory values using continuous regression analysis, adjusting for age, sex, sampling year, and PC1-10 as covariates using the glm function in Plink2. Laboratory values with a significant QTL (FDR < 0.05) were included in follow-up analyses. For mediation analysis^53^, we selected significant labQTLs limiting to the smallest p-value per each KANTA lab value, and included both the SNP and the corresponding KANTA lab value as independent variables in a logistic regression model of blood donorship. We accepted lab value associations reaching FDR < 0.05, indicating that these values associate with blood donorship when adjusted for their QTL SNPs. To further validate the causal effect of each GWAS lead variant on KANTA laboratory values, we employed the Sobel mediation test^54^ using the R package bda v18.3.2, accepting FDR < 0.05.

## Data Availability

FinnGen summary statistics data is available through FinnGen: https://www.finngen.fi/en/access_results

## Data Availability

FinnGen summary statistics and other information are available at https://r12.finngen.fi/. Data supporting the current study are available from the authors upon reasonable request and with permission of FinnGen.

The GWAS summary statistics for the blood donor phenotype will be available at a public repository to be determined later.

## Code Availability

Code will be available in https://github.com/FRCBS/HDE-GWAS.

## Acknowledgements

We want to acknowledge the participants and investigators of FinnGen study. The FinnGen project is funded by two grants from Business Finland (HUS 4685/31/2016 and UH 4386/31/2016) and the following industry partners: AbbVie Inc., AstraZeneca UK Ltd, Biogen MA Inc., Bristol Myers Squibb (and Celgene Corporation & Celgene International II Sàrl), Genentech Inc., Merck Sharp & Dohme LCC, Pfizer Inc., GlaxoSmithKline Intellectual Property Development Ltd., Sanofi US Services Inc., Maze Therapeutics Inc., Janssen Biotech Inc, Novartis Pharma AG, and Boehringer Ingelheim International GmbH. Following biobanks are acknowledged for delivering biobank samples to FinnGen: Auria Biobank (www.auria.fi/biopankki), THL Biobank (www.thl.fi/biobank), Helsinki Biobank (www.helsinginbiopankki.fi), Biobank Borealis of Northern Finland (https://www.ppshp.fi/Tutkimus-ja-opetus/Biopankki/Pages/Biobank-Borealis-briefly-in-English.aspx), Finnish Clinical Biobank Tampere (www.tays.fi/en-US/Research_and_development/Finnish_Clinical_Biobank_Tampere), Biobank of Eastern Finland (www.ita-suomenbiopankki.fi/en), Central Finland Biobank (www.ksshp.fi/fi-FI/Potilaalle/Biopankki), Finnish Red Cross Blood Service Biobank (www.veripalvelu.fi/verenluovutus/biopankkitoiminta), Terveystalo Biobank (www.terveystalo.com/fi/Yritystietoa/Terveystalo-Biopankki/Biopankki/) and Arctic Biobank (https://www.oulu.fi/en/university/faculties-and-units/faculty-medicine/northern-finland-birth-cohorts-and-arctic-biobank). All Finnish Biobanks are members of BBMRI.fi infrastructure (www.bbmri.fi). Finnish Biobank Cooperative -FINBB (https://finbb.fi/) is the coordinator of BBMRI-ERIC operations in Finland. The Finnish biobank data can be accessed through the Fingenious^®^ services (https://site.fingenious.fi/en/) managed by FINBB.

In addition, we want to acknowledge Dr Katri Haimila for her valuable advice with red cell antigen immunogenetics, Dr Kati Hyvärinen for her expertise and kind help with red cell antigen imputation and the personnel of Blood Service Biobank for their kind help and support for the study.

## Author Contributions

Original study concept by JC and JR. Genetic and statistical analysis of the study: JC, JT, MA and JR. Visualization: JC, JT and JR. Medical expertise: JL. FinnGen: data curation and resources. Contributed to the study design: JC, JR, JT, MA, SK, JP. Original manuscript: JC. All the authors read, commented, and approved the final manuscript.

## References

1. Finnish Red Cross Blood Service - donation eligibility. https://www.veripalvelu.fi/en/faq/?search=&category=eligibility.

2. Hu, X. et al. Additive and interaction effects at three amino acid positions in HLA-DQ and HLA-DR molecules drive type 1 diabetes risk. Nat. Genet.47, 898–905 (2015).

3. De Silvestri, A. et al. The Involvement of HLA Class II Alleles in Multiple Sclerosis: A Systematic Review with Meta-analysis. Dis. Markers 2019, 1409069 (2019).

4. Dendrou, C. A., Petersen, J., Rossjohn, J. & Fugger, L. HLA variation and disease. Nat. Rev. Immunol. 18, 325–339 (2018).

5. Ritari, J., Koskela, S., Hyvärinen, K., FinnGen & Partanen, J. HLA-disease association and pleiotropy landscape in over 235,000 Finns. Hum. Immunol. 83, 391–398 (2022).

6. Atsma, F., Veldhuizen, I., Kort, W. De & Vegt, F. De. Healthy donor effect: its magnitude in health research among blood donors. Transfusion 51, 1820–1828 (2011).

7. Marsh, W. L. & Redman, C. M. The Kell blood group system: a review. Transfusion 30, 158–167 (1990).

8. Avent, N. D. & Reid, M. E. The Rh blood group system: a review. Blood 95, 375–387 (2000).

9. Levine, P. & Stetson, R. E. Landmark article July 8, 1939. An unusual case of intra-group agglutination. By Philip Levine and Rufus E Stetson. JAMA 251, 1316–1317 (1984).

10. Giblett, E. R. A critique of the theoretical hazard of inter vs. intra-racial transfusion. Transfusion 1, 233–238 (1961).

11. Stowell, S. R. & Stowell, C. P. Biologic roles of the ABH and Lewis histo-blood group antigens part II: thrombosis, cardiovascular disease and metabolism. Vox Sang. 114, 535–552 (2019).

12. Dahlén, T., Clements, M., Zhao, J., Olsson, M. L. & Edgren, G. An agnostic study of associations between ABO and RhD blood group and phenome-wide disease risk. Elife 10, (2021).

13. Ward, S. E., O’Sullivan, J. M. & O’Donnell, J. S. The relationship between ABO blood group, von Willebrand factor, and primary hemostasis. Blood 136, 2864–2874 (2020).

14. Clancy, J. et al. Blood donor biobank as a resource in personalised biomedical genetic research. Eur. J. Hum. Genet.(2024) doi:10.1038/s41431-023-01528-0.

15. Mitchell, R. Blood banks, biobanks, and the ethics of donation. Transfusion 50, 1866–1869 (2010).

16. Raivola, V., Snell, K., Helén, I. & Partanen, J. Attitudes of blood donors to their sample and data donation for biobanking. Eur. J. Hum. Genet. 27, 1659–1667 (2019).

17. Raivola, V., Snell, K., Helen, I. & Partanen, J. Attitudes of blood donors to their sample and data donation for biobanking. Eur. J. Hum. Genet. 27, 1659–1667 (2019).

18. FinnGen. FinnGen Result Browser. https://r10.finngen.fi/ (2024).

19. National Institutes of Health LDlink. https://ldlink.nih.gov/?tab=home.

20. ISBT Blood Group Allele Tables. https://www.isbtweb.org/isbt-working-parties/rcibgt/blood-group-allele-tables.html.

21. Li, S. & Schooling, C. M. A phenome-wide association study of ABO blood groups. BMC Med. 18, 334 (2020).

22. Bowness, P. HLA-B27. Annu. Rev. Immunol. 33, 29–48 (2015).

23. Simmons, K. M. et al. Failed Genetic Protection: Type 1 Diabetes in the Presence of HLA-DQB1*06:02. Diabetes 69, 1763–1769 (2020).

24. Lazaratos, A.-M., Annis, M. G. & Siegel, P. M. GPNMB: a potent inducer of immunosuppression in cancer. Oncogene 41, 4573–4590 (2022).

25. Szklarczyk, D. et al. The STRING database in 2023: protein-protein association networks and functional enrichment analyses for any sequenced genome of interest. Nucleic Acids Res.51, D638–D646 (2023).

26. Brown, J. H. et al. Three-dimensional structure of the human class II histocompatibility antigen HLA-DR1. Nature 364, 33–39 (1993).

27. Goodin, D. S., Khankhanian, P., Gourraud, P.-A. & Vince, N. Highly conserved extended haplotypes of the major histocompatibility complex and their relationship to multiple sclerosis susceptibility. PLoS One 13, e0190043 (2018).

28. Stowell, C. P. & Stowell, S. R. Biologic roles of the ABH and Lewis histo-blood group antigens Part I: infection and immunity. Vox Sang. 114, 426–442 (2019).

29. Sayers, E. W. et al. Database resources of the national center for biotechnology information. Nucleic Acids Res.50, D20–D26 (2022).

30. Daniels, G. The molecular genetics of blood group polymorphism. Hum. Genet. 126, 729–742 (2009).

31. Che Yaacob, N. S., Islam, M. A., Alsaleh, H., Ibrahim, I. K. & Hassan, R. Alpha-hemoglobin-stabilizing protein (AHSP): a modulatory factor in β-thalassemia. Int. J. Hematol. 111, 352–359 (2020).

32. dos Santos, C. O. & Costa, F. F. AHSP and beta-thalassemia: a possible genetic modifier. Hematology 10, 157–161 (2005).

33. Giardine, B. et al. Updates of the HbVar database of human hemoglobin variants and thalassemia mutations. Nucleic Acids Res. 42, D1063–9 (2014).

34. Rutherford-Parker, N. J., Colby, J. M., Gehrie, E. A. & Booth, G. S. Unrecognized Hemoglobin Variants in the Donor Blood Supply Are Detectable in the Transfused Population. Am. J. Clin. Pathol. 154, 494–498 (2020).

35. Bonacina, F., Pirillo, A., Catapano, A. L. & Norata, G. D. HDL in Immune-Inflammatory Responses: Implications beyond Cardiovascular Diseases. Cells 10, (2021).

36. Lobier, M. et al. The effect of donation activity dwarfs the effect of lifestyle, diet and targeted iron supplementation on blood donor iron stores. PLoS One 14, e0220862 (2019).

37. Toivonen, J., Allara, E., Castrén, J., di Angelantonio, E. & Arvas, M. The value of genetic data from 665,460 individuals in managing iron deficiency anaemia and suitability to donate blood. Vox Sang. 119, 34–42 (2024).

38. Lee H, Aronson JK, N. D. Catalogue of bias collaboration. Collider bias. In Catalogue Of Bias. https://catalogofbias.org/biases/collider-bias/ (2019).

39. Tönnies, T., Kahl, S. & Kuss, O. Collider Bias in Observational Studies. Dtsch. Arztebl. Int. 119, 107–122 (2022).

40. Digitale, J. C., Martin, J. N., Glidden, D. V & Glymour, M. M. Key concepts in clinical epidemiology: collider-conditioning bias. J. Clin. Epidemiol. 161, 152–156 (2023).

41. R Core Team. https://www.r-project.org/.

42. R studio. https://www.rstudio.com/.

43. Benjamini, Y. & Yekutieli, D. The control of the false discovery rate in multiple testing under dependency. Ann. Stat. 29, 1165–1188 (2001).

44. Mbatchou, J. et al. Computationally efficient whole-genome regression for quantitative and binary traits. Nat. Genet. 53, 1097–1103 (2021).

45. Kurki, M. I. et al. FinnGen provides genetic insights from a well-phenotyped isolated population. Nature 613, 508–518 (2023).

46. Wang, G., Sarkar, A., Carbonetto, P. & Stephens, M. A Simple New Approach to Variable Selection in Regression, with Application to Genetic Fine Mapping. J. R. Stat. Soc. Ser. B Stat. Methodol. 82, 1273–1300 (2020).

47. FinnGen. FinnGen Documentation of data release. https://finngen.gitbook.io/documentation/ (2024).

48. Bulik-Sullivan, B. K. et al. LD Score regression distinguishes confounding from polygenicity in genome-wide association studies. Nat. Genet. 47, 291–295 (2015).

49. Zheng, X. et al. HIBAG - HLA genotype imputation with attribute bagging. Pharmacogenomics J. 14, 192–200 (2014).

50. Ritari, J. et al. Increasing accuracy of HLA imputation by a population-specific reference panel in a FinnGen biobank cohort. NAR Genomics Bioinforma. 2, (2020).

51. Hyvärinen, K. et al. A machine-learning method for biobank-scale genetic prediction of blood group antigens. PLoS Comput. Biol. 20, e1011977 (2024).

52. Suhre, K., McCarthy, M. I. & Schwenk, J. M. Genetics meets proteomics: perspectives for large population-based studies. Nat. Rev. Genet. 22, 19–37 (2021).

53. VanderWeele, T. J. Mediation Analysis: A Practitioner’s Guide. Annu. Rev. Public Health 37, 17–32 (2016).

54. Preacher, K. J. & Hayes, A. F. Asymptotic and resampling strategies for assessing and comparing indirect effects in multiple mediator models. Behav. Res. Methods 40, 879–891 (2008).

